# Prognostic accuracy of emergency department triage tools for adults with suspected COVID-19: The PRIEST observational cohort study

**DOI:** 10.1101/2020.09.02.20185892

**Authors:** Ben Thomas, Steve Goodacre, Ellen Lee, Laura Sutton, Amanda Loban, Simon Waterhouse, Richard Simmonds, Katie Biggs, Carl Marincowitz, Jose Schutter, Sarah Connelly, Elena Sheldon, Jamie Hall, Emma Young, Andrew Bentley, Kirsty Challen, Chris Fitzimmons, Tim Harris, Fiona Lecky, Andrew Lee, Ian Maconochie, Darren Walter

## Abstract

**Objectives:** The World Health Organisation (WHO) and National Institute for Health and Care Excellence (NICE) recommend various triage tools to assist decision-making for patients with suspected COVID-19. We aimed to estimate the accuracy of triage tools for predicting severe illness in adults presenting to the emergency department (ED) with suspected COVID-19 infection.

**Methods:** We undertook a mixed prospective and retrospective observational cohort study in 70 EDs across the United Kingdom (UK). We collected data from people attending with suspected COVID-19 between 26 March 2020 and 28 May 2020, and used presenting data to determine the results of assessment with the following triage tools: the WHO algorithm, NEWS2, CURB-65, CRB-65, PMEWS and the swine flu adult hospital pathway (SFAHP). We used 30-day outcome data (death or receipt of respiratory, cardiovascular or renal support) to determine prognostic accuracy for adverse outcome.

**Results:** We analysed data from 20892 adults, of whom 4672 (22.4%) died or received organ support (primary outcome), with 2058 (9.9%) receiving organ support and 2614 (12.5%) dying without organ support (secondary outcomes). C-statistics for the primary outcome were: CURB-65 0.75; CRB-65 0.70; PMEWS 0.77; NEWS2 (score) 0.77; NEWS2 (rule) 0.69; SFAHP (6-point) 0.70; SFAHP (7-point) 0.68; WHO algorithm 0.61. All triage tools showed worse prediction for receipt of organ support and better prediction for death without organ support.

At the recommended threshold, PMEWS and the WHO criteria showed good sensitivity (0.96 and 0.95 respectively), at the expense of specificity (0.31 and 0.27 respectively). NEWS2 showed similar sensitivity (0.96) and specificity (0.28) when a lower threshold than recommended was used.

**Conclusion:** CURB-65, PMEWS and NEWS2 provide good but not excellent prediction for adverse outcome in suspected COVID-19, and predicted death without organ support better than receipt of organ support. PMEWS, the WHO criteria and NEWS2 (using a lower threshold than usually recommended) provide good sensitivity at the expense of specificity.

**Registration:** ISRCTN registry, ISRCTN28342533, http://www.isrctn.com/ISRCTN28342533

## Introduction

The emergency department (ED) has a crucial role in the management of patients with suspected COVID-19. ED management involves assessing the risk of adverse outcome and the need for life-saving intervention, and then using this to determine decisions around admission to hospital and inpatient referral [1,2]. Triage tools can assist decision-making by combining information from clinical assessment in a structured manner to predict the risk of adverse outcome. Triage tools may take the form of a score, which allocates points to risk predictors to indicate an increasing risk of adverse outcome, or a rule, which uses risk predictors to determine a clinical decision, such as hospital admission or discharge. Adults and children presenting to the ED with suspected COVID-19 differ markedly in their need for hospital admission and risk of adverse outcome [3], so they require different triage tools. We focus on adults in this study.

Guidelines have recommended a number of triage tools for adults with suspected COVID-19. The World Health Organisation (WHO) decision-making algorithm for acute respiratory infection [4] recommends hospital admission for severe pneumonia (respiratory rate >30/minute, oxygen saturation <90% or signs of respiratory distress) or respiratory infection associated with comorbidities (age >60, hypertension, diabetes, cardiovascular disease, chronic respiratory disease, chronic renal disease or immunocompromising conditions). The United Kingdom (UK) National Institute for Health and Care Excellence (NICE) COVID-19 rapid guideline [5] suggests that the National Early Warning Score version 2 (NEWS2) score [6] can be useful for predicting the risk of deterioration. NEWS2 uses heart rate, respiratory rate, systolic blood pressure, oxygen saturation, temperature, and conscious level to allocate a score between zero and 20. The guideline also notes that the CRB-65 tool can determine the need for hospital admission in adults with pneumonia but has not been validated in people with COVID-19. The CURB-65 pneumonia score [7] uses five variables (confusion, urea level, respiratory rate, blood pressure, and age) to generate a score between zero and five. The CRB-65 score allows use without blood testing by dropping urea measurement from the score.

Triage tools developed or recommended for an influenza pandemic could be used for suspected COVID-19. Guidance during the 2009 H1N1 pandemic included a swine flu hospital pathway for ED management with seven criteria, any one of which predicts increased risk and the need for hospital assessment [8]. The Pandemic Modified Early Warning Score (PMEWS) [9] uses physiological variables, age, social factors, chronic disease, and performance status to generate a score between zero and nineteen.

Triage tools need to be validated in the clinically relevant population to determine whether they accurately predict adverse outcome. We developed the Pandemic Influenza Triage in the Emergency Department (PAINTED) study following the 2009 H1N1 pandemic to evaluate triage tools for suspected pandemic influenza. We modified the PAINTED protocol to become the Pandemic Respiratory Infection Emergency System Triage (PRIEST) study in January 2020 to address any pandemic respiratory infection and include triage tools recommended for COVID-19.

## Aims and objectives

We aimed to estimate the accuracy of triage tools recommended for predicting severe illness in adults presenting to the ED with suspected COVID-19 infection.

## Methods

### Design

We undertook an observational study to collect standardised predictor variables recorded in the ED, which we then used to evaluate triage tools for predicting adverse outcome up to 30 days after initial hospital presentation. The study did not involve any change to patient care. Hospital admission and discharge decisions were made according to usual practice, informed by local and national guidance.

### Setting and population

We identified consecutive patients presenting to the ED of participating hospitals with suspected COVID-19 infection. Patients were eligible if they met the clinical diagnostic criteria [10] of fever (≥ 37.8°C) and acute onset of persistent cough (with or without sputum), hoarseness, nasal discharge or congestion, shortness of breath, sore throat, wheezing, sneezing. This was determined on the basis of the assessing clinician recording that the patient had suspected COVID-19 or completing a standardised assessment form designed for suspected pandemic respiratory infection [11].

### Interventions

We planned to evaluate triage tools recommended for use in the COVID-19 pandemic or the 2009 H1N1 influenza pandemic, as outlined in the introduction: the WHO algorithm, NEWS2, CURB-65, CRB-65, PMEWS and the swine flu adult hospital pathway (SFAHP). The triage tools are described in Appendix 1. NEWS2 can be used as a score, with thresholds between zero and 20 on the total score, or a rule, with a single threshold of a total score greater than four or a score of three on any parameter. We therefore evaluated the performance of NEWS2 as both a score and a rule. The SFAHP has a criterion (G) that is positive if there is any clinical concern. This is difficult to judge objectively or identify from clinical records, so we evaluated the pathway in two ways: (1) A 6-point rule that did not include parameter G; (2) A 7-point rule in which parameter G was positive if the NEWS2 rule was positive. NEWS2 is widely used in the UK health service to identify clinical concern.

### Data collection

Data collection was both prospective and retrospective. We provided participating EDs with a standardised data collection form that included the predictor variables used in the triage tools [11]. Participating sites could adapt the form to their local circumstances, including integrating it into electronic or paper clinical records to facilitate prospective data collection, or using it as a template for research staff to retrospectively extract data from clinical records. We did not seek consent to collect data but information about the study was provided in the ED and patients could withdraw their data at their request. Patients with multiple presentations to hospital were only included once, using data from the first presentation identified by research staff.

### Outcome measurement

Research staff at participating hospitals reviewed patient records at 30 days after initial attendance to identify any adverse outcomes. Patients who died or required respiratory, cardiovascular or renal support were classified as having an adverse outcome. Patients who survived to 30 days without requiring respiratory, cardiovascular or renal support were classified as having no adverse outcome. Respiratory support was defined as any intervention to protect the patient’s airway or assist their ventilation, including non-invasive ventilation or acute administration of continuous positive airway pressure. It did not include supplemental oxygen alone or nebulised bronchodilators. Cardiovascular support was defined as any intervention to maintain organ perfusion, such as inotropic drugs, or invasively monitor cardiovascular, status, such as central venous pressure or pulmonary artery pressure monitoring, or arterial blood pressure monitoring. It did not include peripheral intravenous cannulation or fluid administration. Renal support was defined as any intervention to assist renal function, such as haemofiltration, haemodialysis or peritoneal dialysis. It did not include intravenous fluid administration.

### Analysis

The primary outcome was death, or respiratory, cardiovascular or renal support, as defined above. We also planned secondary analyses using the following outcomes: (1) Respiratory, cardiovascular, or renal support, to predict need for life-saving treatment; (2) Death without respiratory, cardiovascular, or renal support, to predict poor prognosis. If triage tools are used to determine treatment decisions, such as referral to critical care, then it is helpful to know how well they predict need for treatment rather than a potentially irremediable poor prognosis.

We retrospectively applied each triage tools to the data, excluding pregnant women from analysis of NEWS2. Appendix 1 provides details of scoring and handling missing data for the triage tools. For each tool we plotted the receiver-operating characteristic (ROC) curve and calculated the area under the ROC curve (c-statistic) for discriminating between cases with and without adverse outcome. We calculated sensitivity, specificity, positive predictive value and negative predictive value at the following pre-specified decision making thresholds, based on recommended or usual use: 0-1 versus 2-5 for CURB-65; 0-2 versus 3+ for PMEWS; 0-4 versus 5-20 for the NEWS2 score. The WHO algorithm and Swine Flu Hospital Pathway are positive if any criterion is positive. We used STATA (version 16) for analyses [12].

The sample size was dependent on the size and severity of the pandemic, but based on a previous study in the 2009 H1N1 influenza pandemic we estimated we would need to collect data from 20,000 patients across 40-50 hospitals to identify 200 with an adverse outcome. In the event, the adverse outcome rate in adults was much higher in the COVID-19 pandemic, giving us adequate power to undertake primary and secondary analyses.

### Patient and public involvement

The Sheffield Emergency Care Forum (SECF) is a public representative group interested in emergency care research. [13] Members of SECF advised on the development of the PRIEST study and two members joined the Study Steering Committee. Patients were not involved in the recruitment to and conduct of the study. We are unable to disseminate the findings to study participants directly.

## Results

The PRIEST study recruited 22485 patients from 70 EDs across 53 sites between 26 March 2020 and 28 May 2020. We included 20892 in the analysis after excluding 39 who requested withdrawal of their data, 1530 children, 7 with missing age and 17 with missing outcome data.

Table 1 shows the characteristics of adults in the cohort. Some 13997 (67.0%) were admitted after ED assessment and 6521 (31.2%) ultimately tested positive for COVID-19. Overall, 4672 (22.4%) died or received organ support (primary outcome), with 2058 (9.9%) receiving organ support and 2614 (12.5%) dying without organ support (secondary outcomes). Organ support involved respiratory support for 1944 (9.3%), cardiovascular for 517 (2.5%) and renal support for 218 (1%).

**Table 1:** Characteristics of the study population.

| Characteristic | Statistic/level |  |
| --- | --- | --- |
| Age (years) | N | 20892 |
|  | Mean (SD) | 62.4 (19.7) |
|  | Median (IQR) | 64 (48,79) |
| Sex | Missing | 193 |
|  | Male | 10202 (49.3%) |
|  | Female | 10497 (50.7%) |
| Ethnicity | Missing/prefer not to say | 4199 |
|  | UK/Irish/other white | 14243 (85.3%) |
|  | Asian | 1044 (6.3%) |
|  | Black/African/Caribbean | 640 (3.8%) |
|  | Mixed/multiple ethnic groups | 247 (1.5%) |
|  | Other | 519 (3.1%) |
| Presenting features | Cough | 12986 (62.2%) |
|  | Shortness of breath | 15570 (74.5%) |
|  | Fever | 10276 (49.2%) |
| Symptom duration (days) | N | 18877 |
|  | Mean (SD) | 7.9 (8.9) |
|  | Median (IQR) | 5 (2,10) |
| Heart rate (beats/min) | N | 20461 |
|  | Mean (SD) | 94.9 (21.6) |
|  | Median (IQR) | 93 (80,108) |
| Respiratory rate (breaths/min) | N | 20347 |
|  | Mean (SD) | 23.3 (7) |
|  | Median (IQR) | 22 (18,26) |
| Systolic BP (mmHg) | N | 20299 |
|  | Mean (SD) | 134.6 (24.9) |
|  | Median (IQR) | 133 (118,149) |
| Diastolic BP (mmHg) | N | 20213 |
|  | Mean (SD) | 78.2 (16.1) |
|  | Median (IQR) | 78 (68,88) |
| Temperature (°C) | N | 20232 |
|  | Mean (SD) | 37.1 (1.1) |
|  | Median (IQR) | 37 (36.4,37.8) |
| Oxygen saturation (%) | N | 20633 |
|  | Mean (SD) | 94.7 (6.8) |
|  | Median (IQR) | 96 (94,98) |
| Glasgow Coma Scale | N | 15429 |
|  | Mean (SD) | 14.6 (1.4) |
|  | Median (IQR) | 15 (15,15) |
| AVPU | Missing | 2387 |
|  | Alert | 17569 (94.9%) |
|  | Verbal | 640 (3.5%) |
|  | Pain | 183 (1%) |
|  | Unresponsive | 113 (0.6%) |
| Comorbidities | Hypertension | 6435 (30.8%) |
|  | Heart Disease | 4700 (22.5%) |
|  | Diabetes | 4129 (19.8%) |
|  | Other chronic lung disease | 3764 (18%) |
|  | Asthma | 3408 (16.3%) |
|  | Renal impairment | 1930 (9.2%) |
|  | Active malignancy | 1120 (5.4%) |
|  | Steroid therapy | 557 (2.7%) |
|  | No chronic disease | 5791 (27.7%) |
|  | Hypertension | 6435 (30.8%) |
| Performance status | Missing | 1079 |
|  | Unrestricted normal activity | 10536 (53.2%) |
|  | Limited strenuous activity, can do light | 2371 (12%) |
|  | Limited activity, can self care | 2776 (14%) |
|  | Limited self care | 2649 (13.4%) |
|  | Bed/chair bound, no self care | 1481 (7.5%) |
| Other clinical concern | Severe respiratory distress | 587 (2.8%) |
|  | Respiratory exhaustion | 292 (1.4%) |
|  | Severe dehydration | 261 (1.2%) |

Table 2 shows the results for the primary analysis, Table 3 the results for secondary analysis predicting receipt of organ support and Table 4 the results for secondary analysis predicting death without organ support. The ROC curves for these analyses are shown in Figures 1 to 3.

**Table 2.**
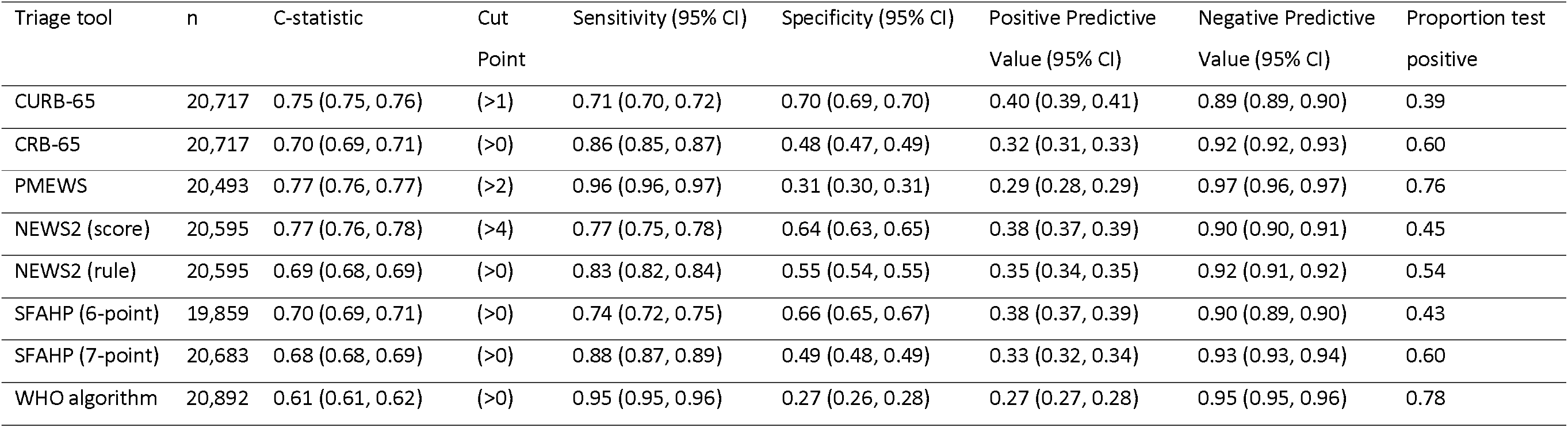
Summary of ROC analysis for existing triage tools predicting adverse outcomes.

| Triage tool | n | C-statistic | Cut Point | Sensitivity (95% CI) | Specificity (95% CI) | Positive Predictive Value (95% CI) | Negative Predictive Value (95% CI) | Proportion test positive |
| --- | --- | --- | --- | --- | --- | --- | --- | --- |
| CURB-65 | 20,717 | 0.75 (0.75, 0.76) | (>1) | 0.71 (0.70, 0.72) | 0.70 (0.69, 0.70) | 0.40 (0.39, 0.41) | 0.89 (0.89, 0.90) | 0.39 |
| CRB-65 | 20,717 | 0.70 (0.69, 0.71) | (>0) | 0.86 (0.85, 0.87) | 0.48 (0.47, 0.49) | 0.32 (0.31, 0.33) | 0.92 (0.92, 0.93) | 0.60 |
| PMEWS | 20,493 | 0.77 (0.76, 0.77) | (>2) | 0.96 (0.96, 0.97) | 0.31 (0.30, 0.31) | 0.29 (0.28, 0.29) | 0.97 (0.96, 0.97) | 0.76 |
| NEWS2 (score) | 20,595 | 0.77 (0.76, 0.78) | (>4) | 0.77 (0.75, 0.78) | 0.64 (0.63, 0.65) | 0.38 (0.37, 0.39) | 0.90 (0.90, 0.91) | 0.45 |
| NEWS2 (rule) | 20,595 | 0.69 (0.68, 0.69) | (>0) | 0.83 (0.82, 0.84) | 0.55 (0.54, 0.55) | 0.35 (0.34, 0.35) | 0.92 (0.91, 0.92) | 0.54 |
| SFAHP (6-point) | 19,859 | 0.70 (0.69, 0.71) | (>0) | 0.74 (0.72, 0.75) | 0.66 (0.65, 0.67) | 0.38 (0.37, 0.39) | 0.90 (0.89, 0.90) | 0.43 |
| SFAHP (7-point) | 20,683 | 0.68 (0.68, 0.69) | (>0) | 0.88 (0.87, 0.89) | 0.49 (0.48, 0.49) | 0.33 (0.32, 0.34) | 0.93 (0.93, 0.94) | 0.60 |
| WHO algorithm | 20,892 | 0.61 (0.61, 0.62) | (>0) | 0.95 (0.95, 0.96) | 0.27 (0.26, 0.28) | 0.27 (0.27, 0.28) | 0.95 (0.95, 0.96) | 0.78 |

**Table 3.** Summary of ROC analysis for existing triage tools predicting organ support.

| Triage tool | n | C-statistic | Cut Point | Sensitivity (95% CI) | Specificity (95% CI) | Positive Predictive Value (95% CI) | Negative Predictive Value (95% CI) | Proportion test positive |
| --- | --- | --- | --- | --- | --- | --- | --- | --- |
| CURB-65 | 20,717 | 0.60 (0.59, 0.61) | (>1) | 0.52 (0.50, 0.54) | 0.62 (0.61, 0.63) | 0.13 (0.12, 0.14) | 0.92 (0.92, 0.93) | 0.39 |
| CRB-65 | 20,717 | 0.58 (0.56, 0.59) | (>0) | 0.74 (0.72, 0.76) | 0.42 (0.41, 0.42) | 0.12 (0.12, 0.13) | 0.94 (0.93, 0.94) | 0.60 |
| PMEWS | 20,493 | 0.68 (0.67, 0.69) | (>2) | 0.94 (0.93, 0.95) | 0.27 (0.26, 0.27) | 0.12 (0.12, 0.13) | 0.98 (0.97, 0.98) | 0.76 |
| NEWS2 (score) | 20,595 | 0.72 (0.71, 0.73) | (>4) | 0.76 (0.74, 0.78) | 0.58 (0.58, 0.59) | 0.17 (0.16, 0.17) | 0.96 (0.95, 0.96) | 0.45 |
| NEWS2 (rule) | 20,595 | 0.65 (0.64, 0.66) | (>0) | 0.81 (0.79, 0.83) | 0.49 (0.49, 0.50) | 0.15 (0.14, 0.16) | 0.96 (0.96, 0.96) | 0.54 |
| SFAHP (6-point) | 19,859 | 0.64 (0.63, 0.65) | (>0) | 0.68 (0.66, 0.71) | 0.60 (0.59, 0.61) | 0.16 (0.15, 0.17) | 0.95 (0.94, 0.95) | 0.43 |
| SFAHP (7-point) | 20,683 | 0.65 (0.64, 0.65) | (>0) | 0.86 (0.84, 0.87) | 0.43 (0.42, 0.44) | 0.14 (0.14, 0.15) | 0.97 (0.96, 0.97) | 0.60 |
| WHO algorithm | 20,892 | 0.57 (0.57, 0.58) | (>0) | 0.91 (0.90, 0.92) | 0.24 (0.23, 0.24) | 0.12 (0.11, 0.12) | 0.96 (0.96, 0.97) | 0.78 |

**Table 4.**
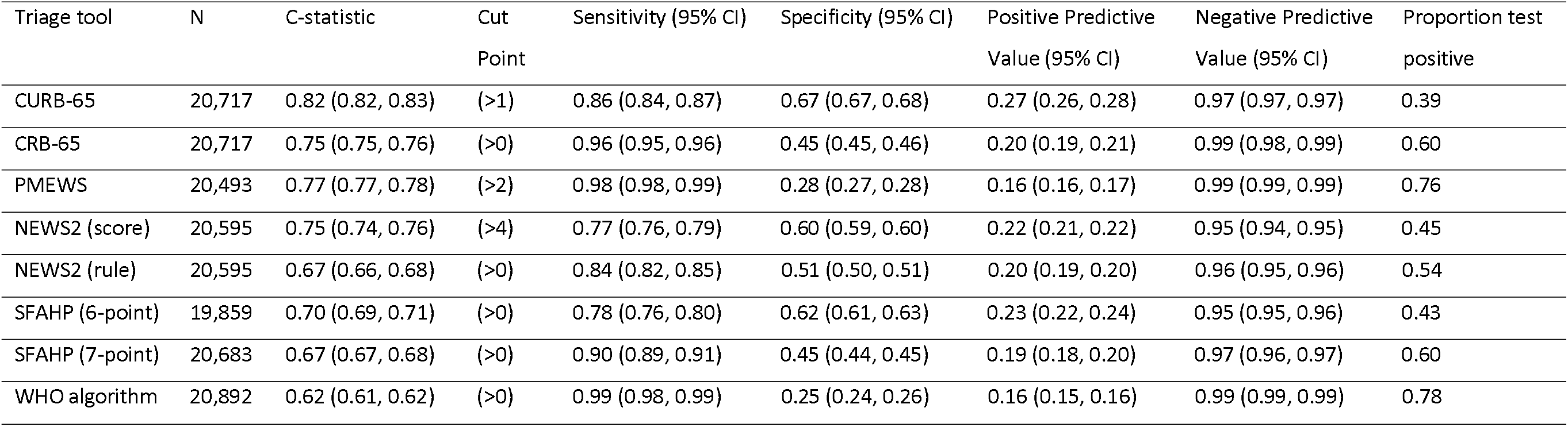
Summary of ROC analysis for existing triage tools predicting death without organ support.

| Triage tool | N | C-statistic | Cut<br>Point | Sensitivity (95% CI) | Specificity (95% CI) | Positive Predictive<br>Value (95% CI) | Negative Predictive<br>Value (95% CI) | Proportion test<br>positive |
| --- | --- | --- | --- | --- | --- | --- | --- | --- |
| CURB-65 | 20,717 | 0.82 (0.82, 0.83) | (>1) | 0.86 (0.84, 0.87) | 0.67 (0.67, 0.68) | 0.27 (0.26, 0.28) | 0.97 (0.97, 0.97) | 0.39 |
| CRB-65 | 20,717 | 0.75 (0.75, 0.76) | (>0) | 0.96 (0.95, 0.96) | 0.45 (0.45, 0.46) | 0.20 (0.19, 0.21) | 0.99 (0.98, 0.99) | 0.60 |
| PMEWS | 20,493 | 0.77 (0.77, 0.78) | (>2) | 0.98 (0.98, 0.99) | 0.28 (0.27, 0.28) | 0.16 (0.16, 0.17) | 0.99 (0.99, 0.99) | 0.76 |
| NEWS2 (score) | 20,595 | 0.75 (0.74, 0.76) | (>4) | 0.77 (0.76, 0.79) | 0.60 (0.59, 0.60) | 0.22 (0.21, 0.22) | 0.95 (0.94, 0.95) | 0.45 |
| NEWS2 (rule) | 20,595 | 0.67 (0.66, 0.68) | (>0) | 0.84 (0.82, 0.85) | 0.51 (0.50, 0.51) | 0.20 (0.19, 0.20) | 0.96 (0.95, 0.96) | 0.54 |
| SFAHP (6-point) | 19,859 | 0.70 (0.69, 0.71) | (>0) | 0.78 (0.76, 0.80) | 0.62 (0.61, 0.63) | 0.23 (0.22, 0.24) | 0.95 (0.95, 0.96) | 0.43 |
| SFAHP (7-point) | 20,683 | 0.67 (0.67, 0.68) | (>0) | 0.90 (0.89, 0.91) | 0.45 (0.44, 0.45) | 0.19 (0.18, 0.20) | 0.97 (0.96, 0.97) | 0.60 |
| WHO algorithm | 20,892 | 0.62 (0.61, 0.62) | (>0) | 0.99 (0.98, 0.99) | 0.25 (0.24, 0.26) | 0.16 (0.15, 0.16) | 0.99 (0.99, 0.99) | 0.78 |

**Figure 1:**
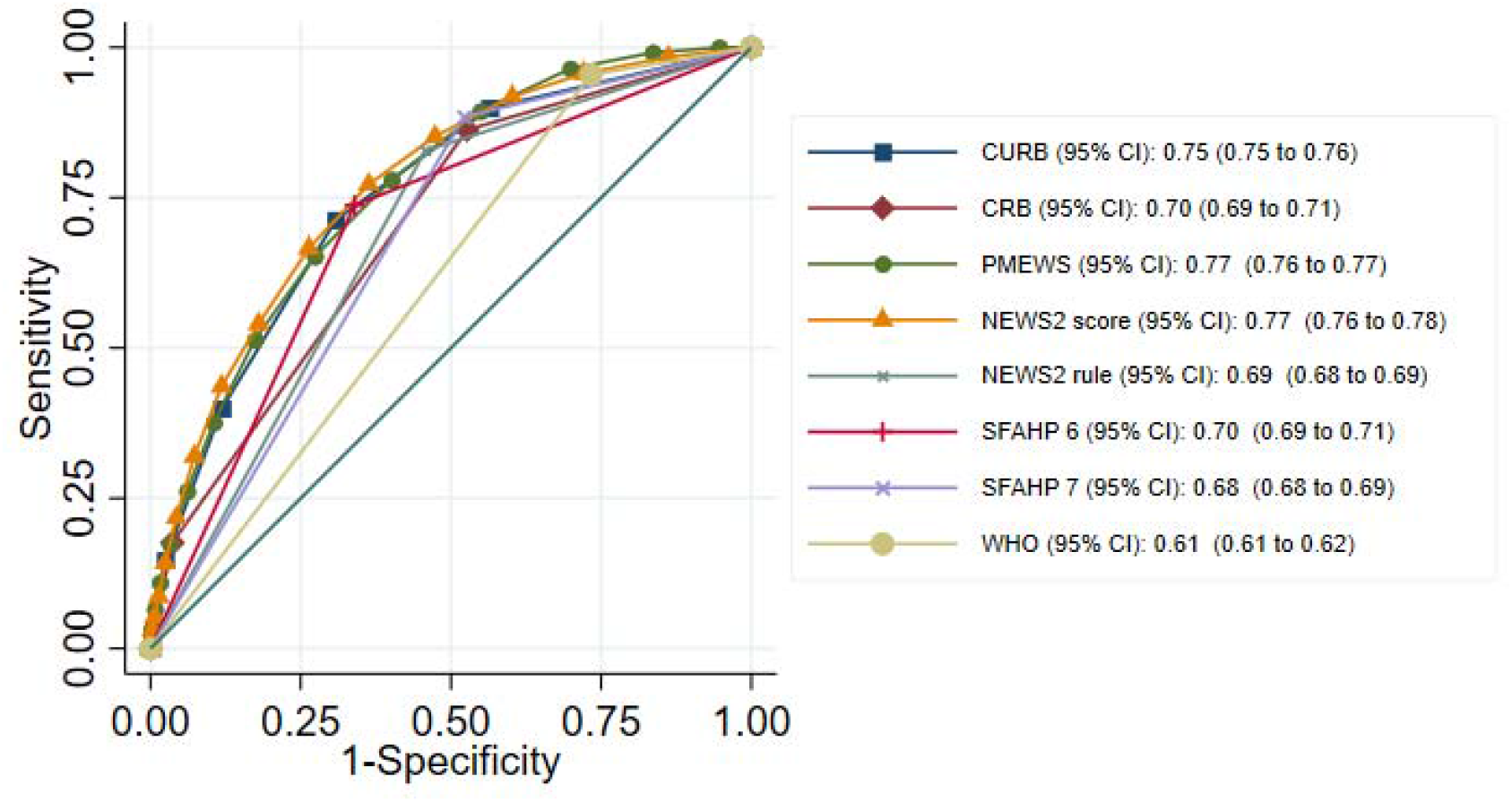
Overlaid ROC curves for triage tools predicting adverse outcome in adults

**Figure 2:**
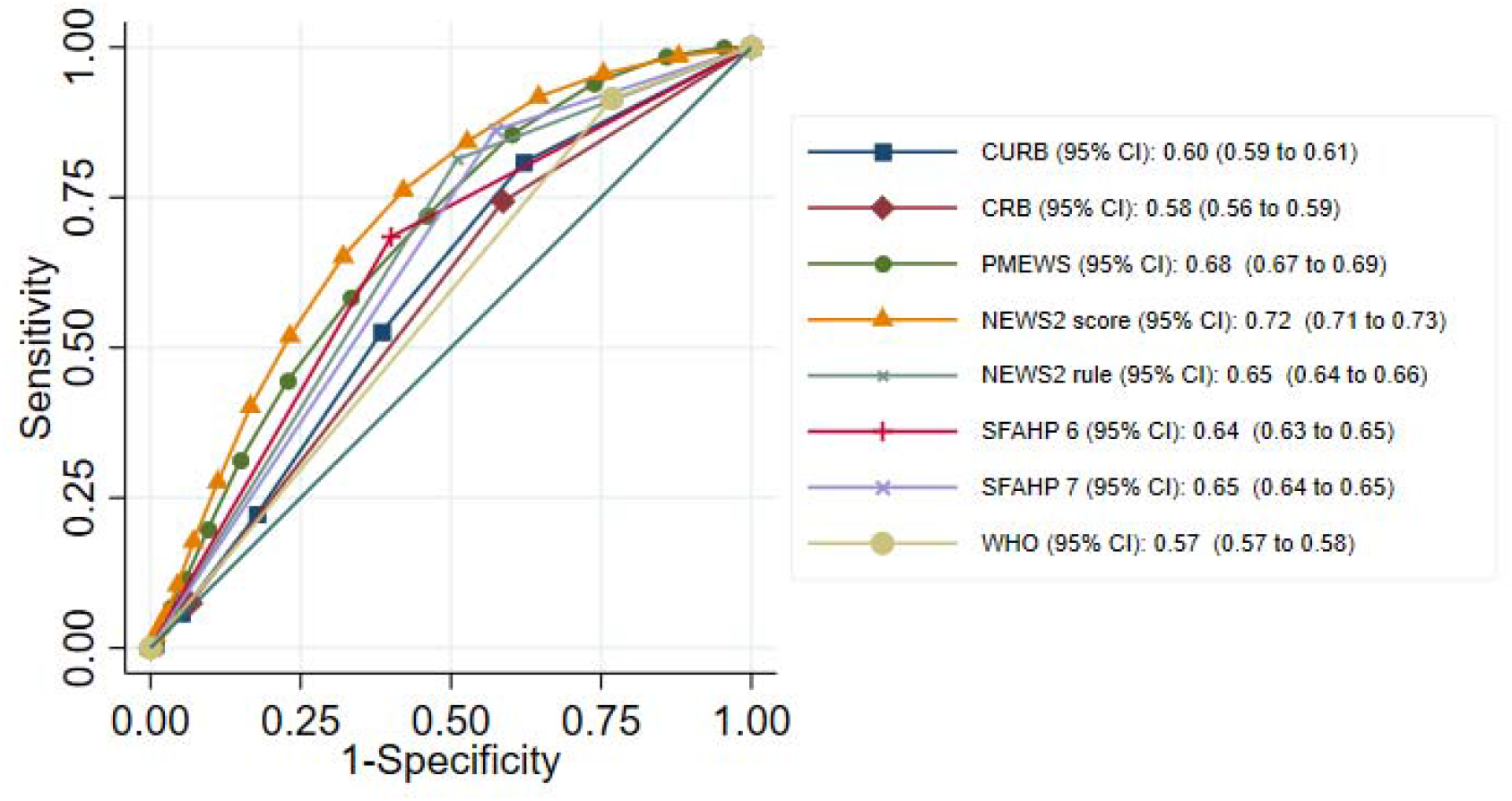
Overlaid ROC curves for triage tools predicting any support in adults

**Figure 3:**
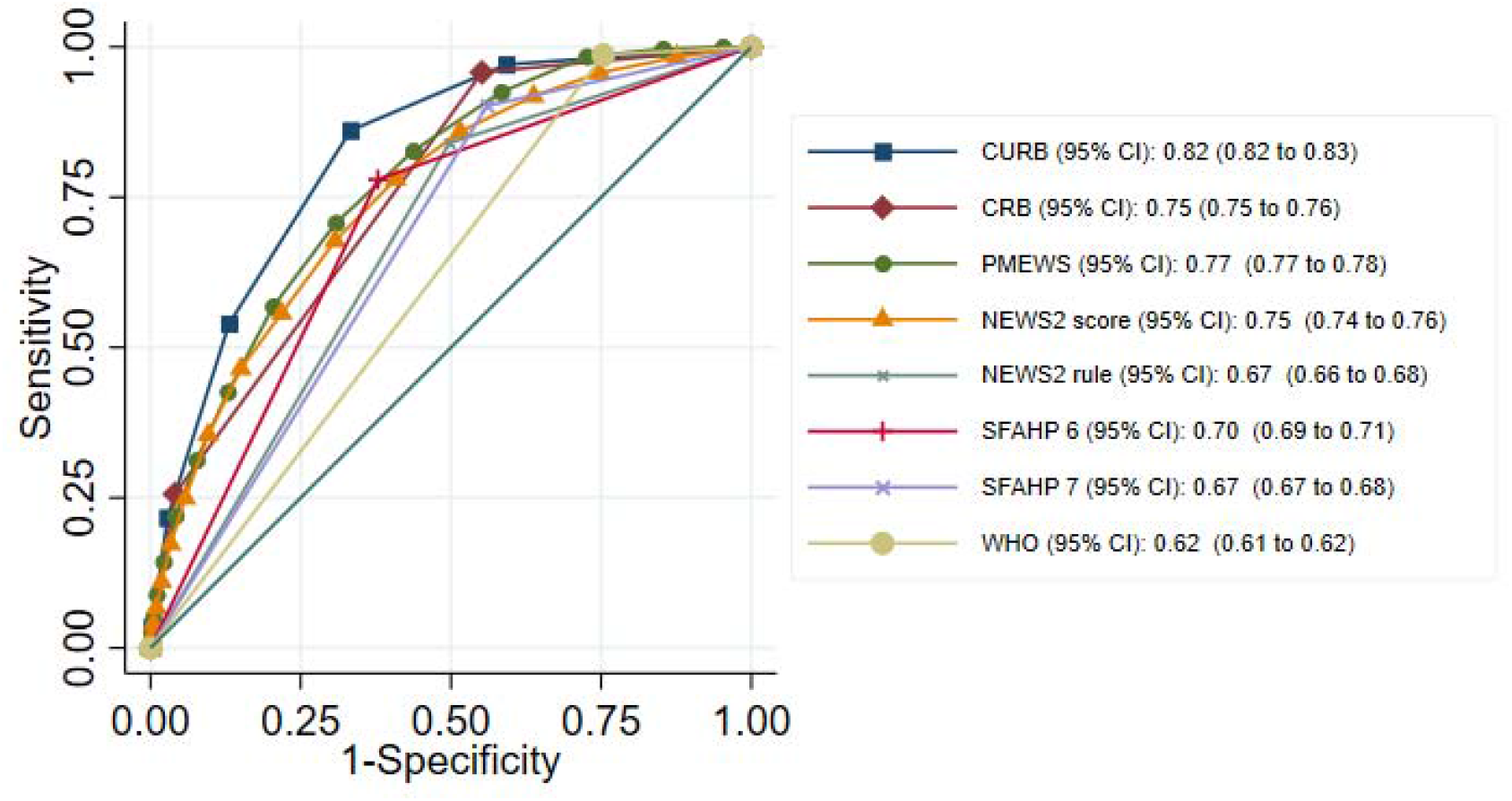
Overlaid ROC curves for triage tools predicting death without support in adults

In the primary analysis, none of the triage tools showed excellent discrimination (c-statistic>0.8) but CURB-65, PMEWS, and NEWS2 showed good discrimination (>0.7). This may reflect the use of multiple points across these tools, as opposed to a single decision-making threshold for other tools. At the pre-specified threshold, PMEWS and the WHO criteria showed good sensitivity (0.96 and 0.95 respectively), at the expense of specificity (0.31 and 0.27 respectively). The sensitivities of other triage tools at the pre-specified threshold were below 0.9, albeit with higher specificities.

The triage tools generally showed worse prediction for receipt of organ support and better prediction for death without organ support. This was most marked for CURB-65 and CRB-65, and least marked for NEWS2. Only NEWS2 showed good prediction for organ support (c-statistic>0.7).

Supplementary Table S1 shows the sensitivity and specificity at each threshold for the triage tools with multiple potential thresholds for decision-making (CURB-65, CRB-65, PMEWS, and NEWS2). These results suggest that NEWS2 could offer good sensitivity (0.96), at the expense of specificity (0.28), if we use a score greater than one to predict adverse outcome. The sensitivity of CURB-65 is 0.90 and CRB-65 is 0.86 at the lowest threshold (any score above zero predicts adverse outcome).

## Discussion

### Summary of main findings

ED clinicians usually use triage tools to support decisions, such as admission to hospital, where sensitivity needs to be optimised at the expense of specificity to avoid missed opportunities to predict and prevent adverse outcome. Our analysis suggests that the WHO algorithm or PMEWS greater than two provide good sensitivity at the expense of specificity, and could be used to support decision-making where sensitivity needs to be optimised. NEWS2 needs to use a lower threshold (any score above one) than currently recommended to achieve a comparable balance of sensitivity and specificity.

The triage tools predicted death without organ support better than they predicted receipt of organ support. Only NEWS2 predicted receipt of organ support with good accuracy. This reflects NEWS2 using only physiological measures, while other triage tools include age, performance status or comorbidities that are more likely to predict death without organ support.

### Previous research

Studies undertaken during the H1N1 influenza 2009 pandemic suggested that existing triage tools have suboptimal accuracy for predicting adverse outcome in acute respiratory infections, with c-statistics below 0.8 [14,-16]. Recent studies have evaluated NEWS2, CURB-65 and CRB-65 in adult inpatients with confirmed COVID-19. Fan *et al* (N=654) [17] reported c-statistics of 0.81, 0.85 and 0.80 respectively for NEWS2, CURB-65 and CRB-65 as predictors of in-hospital death. The conventional thresholds for positivity of scores above four, one and zero offered suboptimal sensitivity (0.79, 0.63 and 0.83), with corresponding specificities of 0.69, 0.91 and 0.69. Bradley et al (N=830) [18] reported c-statistics of 0.67 for NEWS2 and 0.74 for CURB-65 as predictors of 30-day mortality, with sensitivities and specificities at conventional thresholds of 0.83 and 0.37 for NEWS2, and 0.80 and 0.59 for CURB-65. Ma et al (N=305) [19] reported c-statistics of 0.79 for NEWS2 and 0.85 for CURB-65 for predicting death. Satici et al (N=681) [20] reported a c-statistic of 0.79 for predicting 30-day mortality with CURB-65, with sensitivity 0.73 and specificity 0.85 at the conventional threshold. Nguyen et al [21] reported that 36/171 (21%) patients with CURB-65 scores of zero or one died or received intensive care admission. Gidari *et al* (N=68) [22] evaluated NEWS2 as a predictor of intensive care admission and Myrstad et al (N=66) [23] evaluated NEWS2 and CRB65 as predictors of death or intensive care admission, but the small sizes produced imprecise estimates of prognostic parameters.

These studies concur with our findings that the conventional thresholds for NEWS2 and CURB-65 offer inadequate sensitivity to support discharge decisions after ED assessment. The larger studies used 30-day or in-hospital mortality as their outcome. Our analysis suggests that this may overestimate prognostic accuracy if the tools are used to predict need for life-saving treatment rather than simply predicting mortality.

### Strengths and limitations

We collected data from a clinically relevant population of patients presenting with suspected COVID19 across a large and varied range of EDs. The large sample size and high rate of adverse outcome allowed us to estimate parameters with a high degree of precision in primary and secondary analyses. The main limitation is that the triage tools applied to data collected from clinical record review or a standardised data collection form, rather than being applied directly to the patient by the assessing clinician. This may have led to under-estimation of the performance of the triage tool, especially when relevant data were missing. Table 1 shows that data were relatively complete for age, physiological variables and performance status, but the recording of other parameters (respiratory distress, respiratory exhaustion, dehydration) was limited by inability to determine whether the feature was not present or not recorded. This is most salient for the Swine Flu Hospital Pathway and may have led to underestimation of the sensitivity of this triage tool. Another potential limitation is that we may have missed adverse outcomes if patients attended a different hospital after initial hospital discharge. This is arguably less likely in the context of a pandemic, in which movements between regions were curtailed, but cannot be discounted. Finally, although some triage tools can be used in the prehospital or community setting, we recommend caution in extrapolating our findings to other settings, where there may be a lower prevalence of adverse outcome.

### Implications for practice

Our findings suggest that the WHO algorithm or PMEWS greater than two could be used to support hospital admission decisions, providing good sensitivity at the expense of specificity. NEWS2 would need to use a threshold greater than one to achieve a similar balance of sensitivity and specificity. If a triage tool is used to select patients for higher levels of treatment, rather than simply predict risk of adverse outcome, then NEWS2 offers better discrimination than other triage tools. Use of triage tools for this purpose may also require a different balance of sensitivity and specificity, with a higher threshold being used to ensure higher levels of care are reserved for those most likely to benefit.

In general, however, the accuracy of the triage tools evaluated was far from optimal, especially for predicting receipt of organ support. This is arguably unsurprising since they were developed for a variety of purposes and none were derived using data from patients presenting to the ED with suspected COVID-19. Research to derive and validate triage tools specific for COVID-19 is therefore an urgent priority.

### Ethical approval

The North West - Haydock Research Ethics Committee gave a favourable opinion on the PAINTED study on 25 June 2012 (reference 12/NW/0303) and on the updated PRIEST study on 23rd March 2020. The Confidentiality Advisory Group of the Health Research Authority granted approval to collect data without patient consent in line with Section 251 of the National Health Service Act 2006.

## Data Availability

Anonymised data are available from the corresponding author upon reasonable request (contact details on first page). The Confidentiality Advisory Group of the Health Research Authority will need to consider any requests for data to be used for purposes other than those specified in our application, so a data request should be accompanied by explanation of the purpose of the request and justification of the public benefit. We also recommend inclusion of a pre-specified plan of analysis.

## Contributor and guarantor information

SG, AB, KC, CF, TH, FL, ALe, IM and DW conceived and designed the study. BT, KB, ALo, SW, RS, JS, SC, ES, JH and EY acquired the data. EL, LS, SG, BT, KB and CM analysed the data. SG, AB, KC, CF, TH, FL, ALe, IM, DW, EL, LS, SG, BT, KB and CM interpreted the data. All authors contributed to drafting the manuscript. Steve Goodacre is the guarantor of the paper. The corresponding author attests that all listed authors meet authorship criteria and that no others meeting the criteria have been omitted.

## Acknowledgements

We thank Katie Ridsdale for clerical assistance with the study, Erica Wallis (Sponsor representative, all members of the Study Steering Committee (Appendix 2) and the site research teams who delivered the data for the study (Appendix 3), and the research team at the University of Sheffield past and present (Appendix 4).

---

**Supplementary Table S1:**
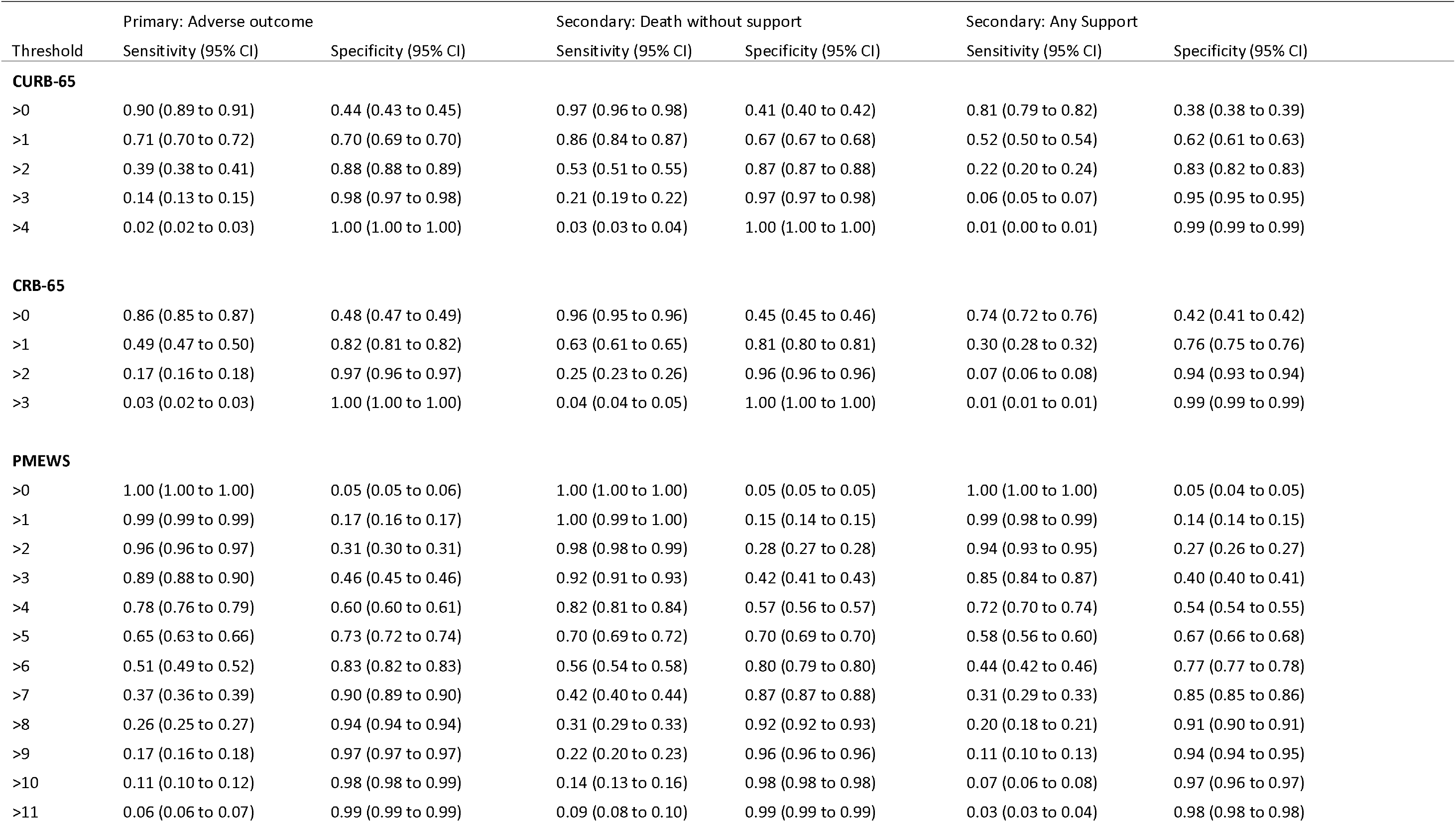
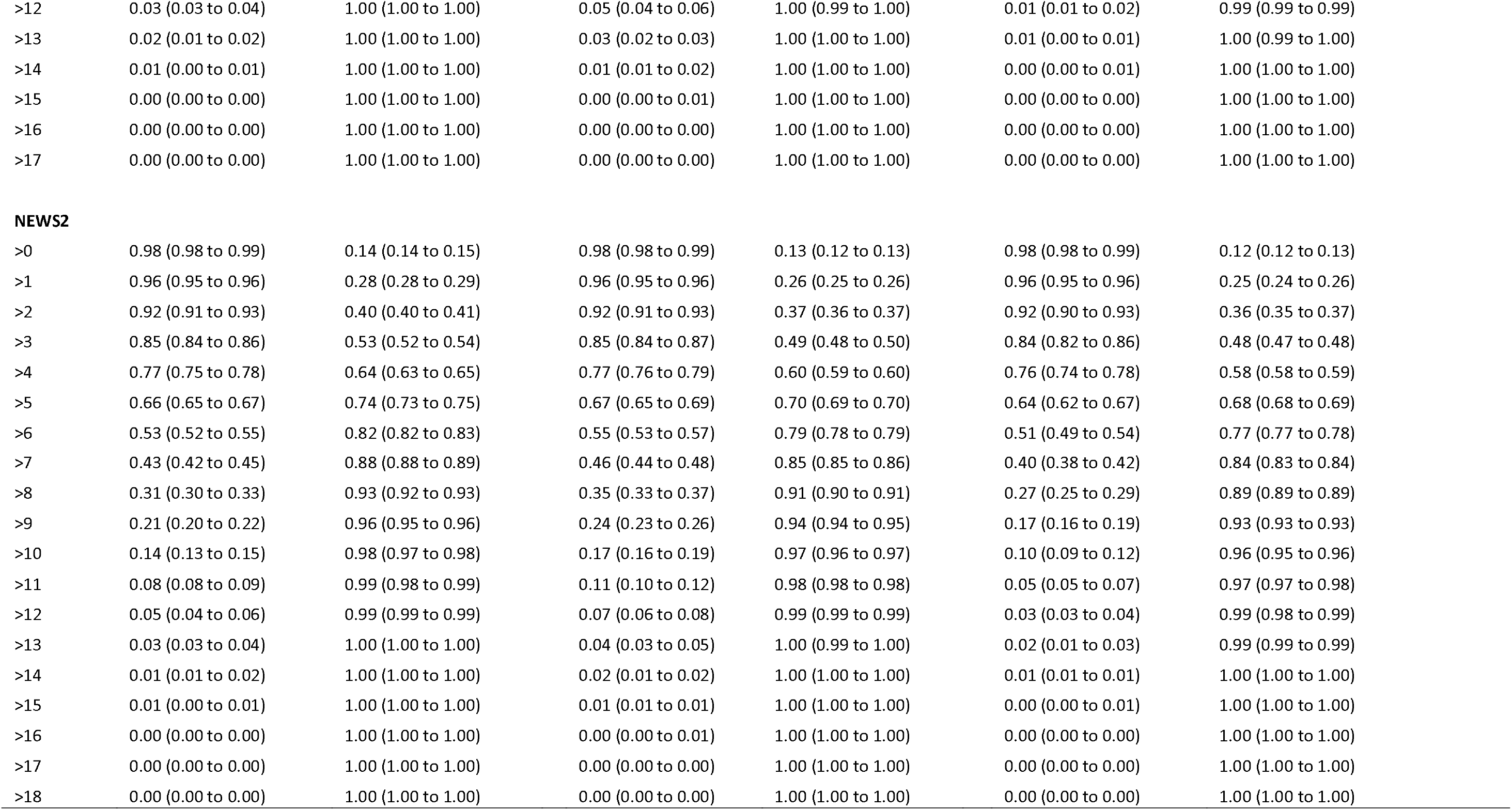
Sensitivity and specificity of each threshold of CURB, CRB, PMEWS and NEWS2 for predicting the primary and secondary outcomes

## Appendix 1: Triage tool scoring details

### CURB-65 and CRB-65

The CURB-65 score uses five parameters, each scoring one point when positive and zero if negative, to give a total score between zero and five. The CRB-65 score does not include urea, so the total score is between zero and four.

Five parameters:

1. Confusion: Glasgow Coma Score (GCS) is less than 15 or AVPU is recorded as anything less than alert (A)
2. Urea: Raised blood urea over 7mmol/litre
3. Respiratory: rate of 30 breaths per minute or more
4. Blood pressure: diastolic BP is 60mmHg or less or systolic BP is 90 mmHg or less
5. Age: 65 years or more

If data for a parameter was missing, we scored the parameter as zero.

### PMEWS

PMEWS uses six physiological and patient parameters to calculate a score from zero to 19. The score is calculated by taking the score in the table below dependent on each of the six physiological parameters then adding points for two patient parameters after if they are positive.

Physiological:

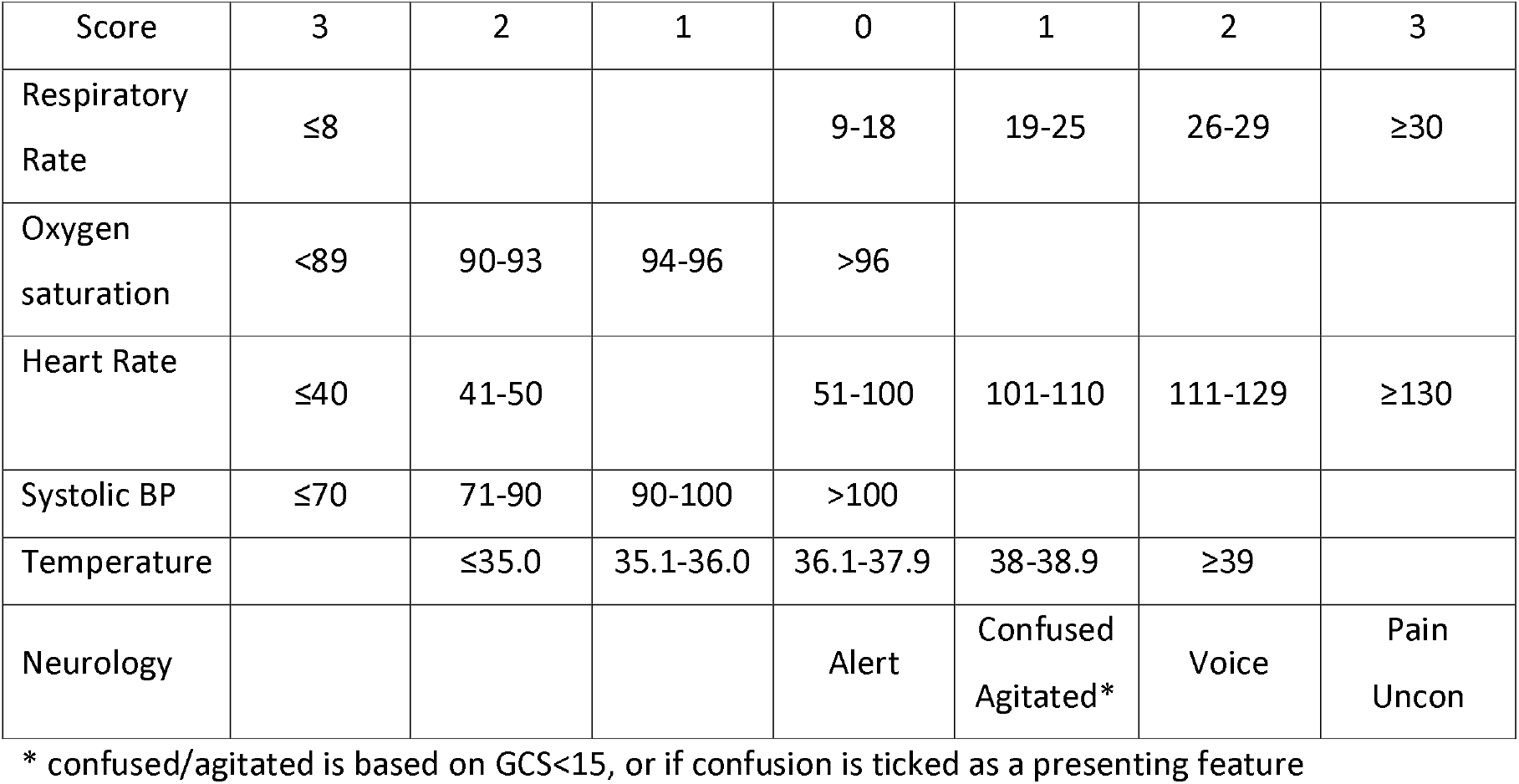

| Score | 3 | 2 | 1 | 0 | 1 | 2 | 3 |
| --- | --- | --- | --- | --- | --- | --- | --- |
| Respiratory Rate | ≤8 |  |  | 9-18 | 19-25 | 26-29 | ≥30 |
| Oxygen saturation | <89 | 90-93 | 94-96 | >96 |  |  |  |
| Heart Rate | ≤40 | 41-50 |  | 51-100 | 101-110 | 111-129 | ≥130 |
| Systolic BP | ≤70 | 71-90 | 90-100 | >100 |  |  |  |
| Temperature |  | ≤35.0 | 35.1-36.0 | 36.1-37.9 | 38-38.9 | ≥39 |  |
| Neurology |  |  |  | Alert | Confused<br>Agitated* | Voice | Pain<br>Uncon |

\* confused/agitated is based on GCS<15, or if confusion is ticked as a presenting feature

Patient:

1. Add 1 point if age>65
2. Add 1 point if either:
  a. Patient lives alone / no fixed abode or
  b. has a co-morbidity (respiratory, cardiac, renal, immunosuppressed, diabetes)
  c. performance status is more than two suggesting limited activity can self-care, limited activity limited self-care, or bed/chair bound no self-care.

If data for a parameter was missing, we scored the parameter as zero. If more than three parameters were missing, we did not calculate a score for the patient. If AVPU was missing and GCS was recorded, we imputed the following AVPU scores using GCS: Alert if GCS=15, Verbal response if GCS=12-14; Pain response if GCS=9-11 and Unconscious if GCS<9.

### Swine Flu Adult Hospital Pathway

The Swine Flu Adult Hospital Pathway consists of seven criteria operating as a rule, with the rule being positive if any criteria reaches its threshold.

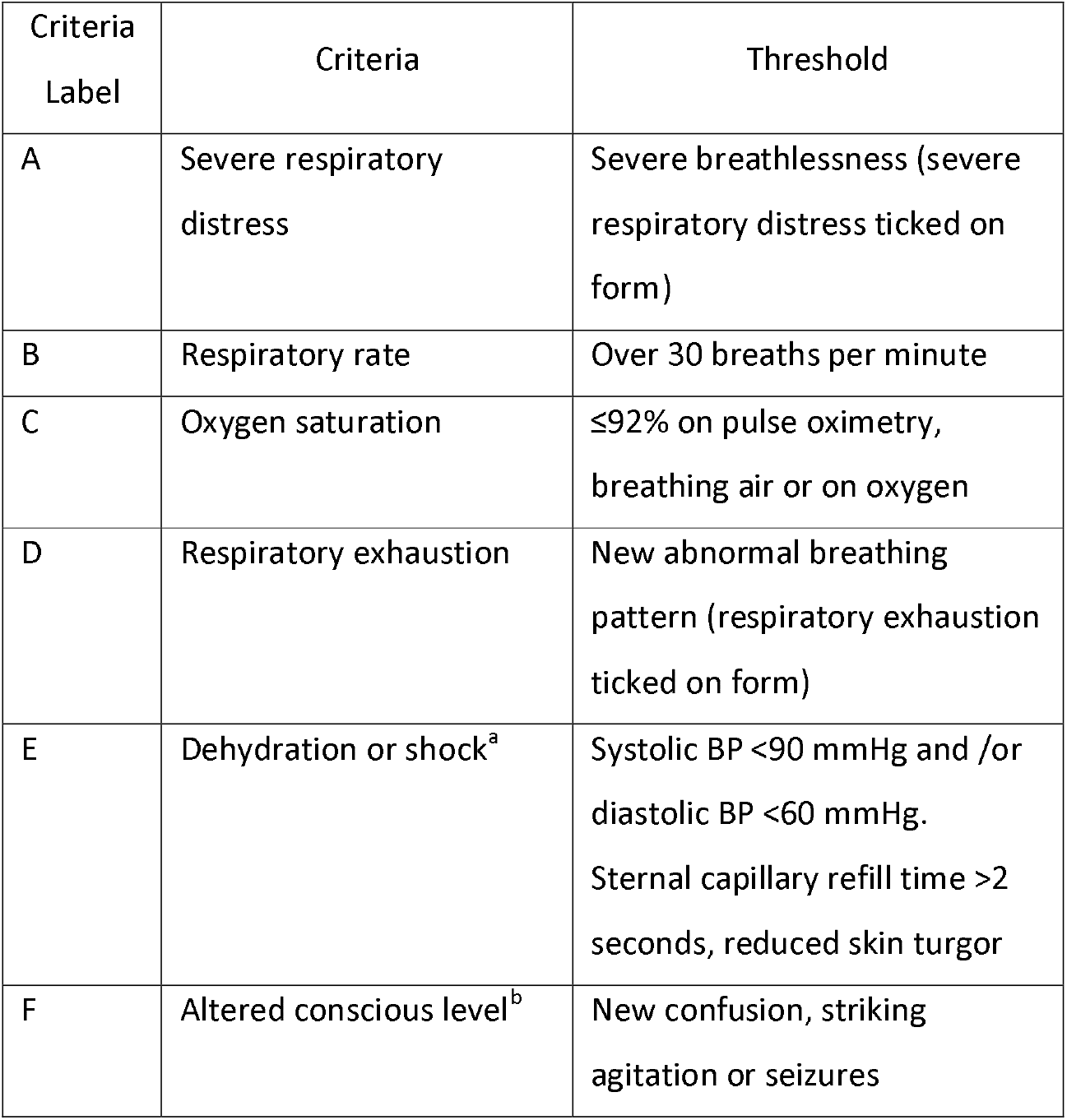
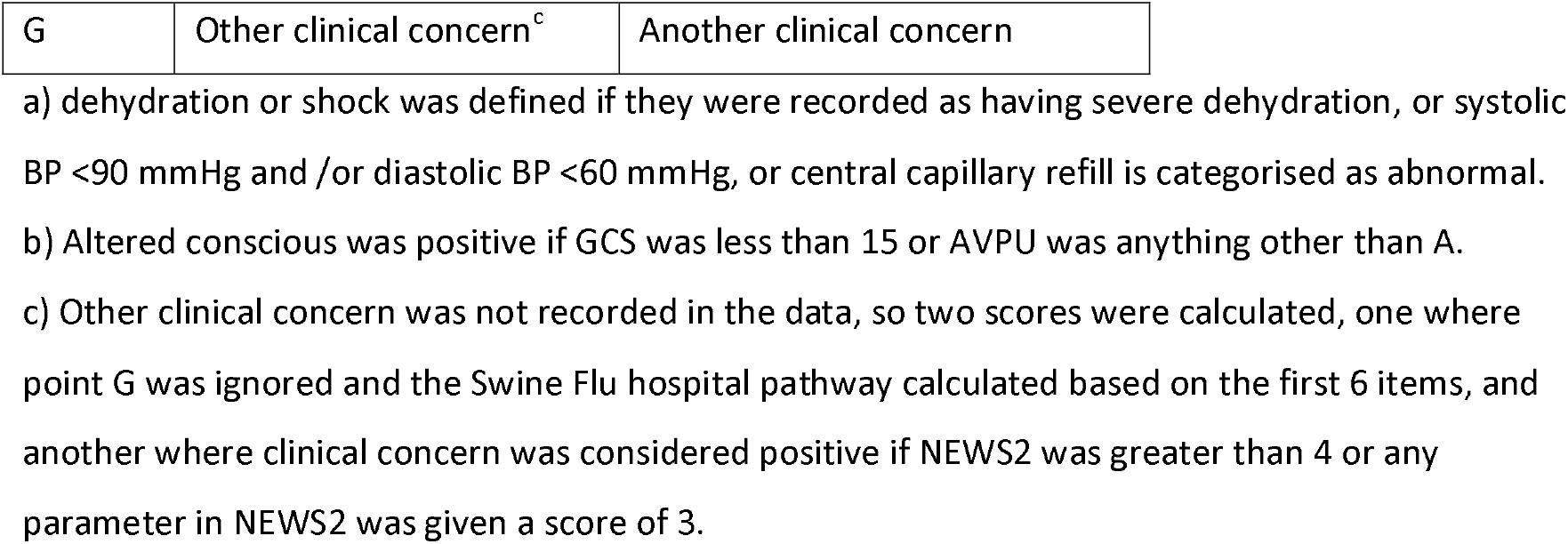

If data for a parameter was missing, we defined the parameter as being negative. If more than three parameters were missing, we did not calculate a score for the patient.

### NEWS2

The NEWS2 has seven parameters, each of which are scores from zero to three providing an overall score between zero and 20. The scores for each parameter can be found in the table below.

| Score | 3 | 2 | 1 | 0 | 1 | 2 | 3 |
| --- | --- | --- | --- | --- | --- | --- | --- |
| Respiratory Rate | ≤8 |  | 9-11 | 12-20 |  | 21-24 | ≥25 |
| Oxygen saturation | ≤91 | 92-93 | 94-95 | ≥96 |  |  |  |
| Heart Rate | ≤40 |  | 41-50 | 51-90 | 91-110 | 111-130 | ≥131 |
| Systolic BP | ≤90 | 91-100 | 101-110 | 111-219 |  |  | ≥220 |
| Temperature | ≤35.0 |  | 35.1-36.0 | 36.1-38.0 | 38.1-39.0 | ≥39.1 |  |
| Neurology |  |  |  | Alert |  |  | Confusion,<br>Voice, Pain,<br>Unresponsive |
| Air or Oxygen |  | Oxygen<br>(based on<br>FiO<sub>2</sub>>21%, or<br>FiO<sub>2</sub>>0<br>L/min) |  | Air |  |  |  |

If data for a parameter was missing, we scored the parameter as zero. If more than three parameters were missing, we did not calculate a score for the patient. The scale for patients with confirmed hypercapnic respiratory failure was not used. We analysed NEWS2 in two ways: (1) As a score, with thresholds between zero and 20 on the total score; (2) As a rule, with a single threshold of a total score greater than four or a score of three on any parameter.

### The WHO decision-making algorithm for hospitalisation with pneumonia

The WHO decision-making algorithm for hospitalisation with pneumonia recommends admission for an adult patient (rule positive) if any of the following are present:

- respiratory rate >30/minute,
- oxygen saturation <90%,
- respiratory distress,
- age >60,
- any of the following comorbidities; hypertension, diabetes, cardiovascular disease, chronic respiratory disease, renal impairment immunosuppression

If data for a parameter was missing, we assumed it was negative. If more than three parameters were missing, we did not calculate a score for the patient

